# The E484K mutation in the SARS-CoV-2 spike protein reduces but does not abolish neutralizing activity of human convalescent and post-vaccination sera

**DOI:** 10.1101/2021.01.26.21250543

**Authors:** Sonia Jangra, Chengjin Ye, Raveen Rathnasinghe, Daniel Stadlbauer, PVI study group, Florian Krammer, Viviana Simon, Luis Martinez-Sobrido, Adolfo García-Sastre, Michael Schotsaert

## Abstract

One year in the coronavirus disease 2019 (COVID-19) pandemic, the first vaccines are being rolled out under emergency use authorizations. It is of great concern that newly emerging variants of severe acute respiratory syndrome coronavirus 2 (SARS-CoV-2) can escape antibody-mediated protection induced by previous infection or vaccination through mutations in the spike protein. The glutamate (E) to Lysine (K) substitution at position 484 (E484K) in the receptor binding domain (RBD) of the spike protein is present in the rapidly spreading variants of concern belonging to the B.1.351 and P.1 lineages. We performed *in vitro* microneutralization assays with both the USA-WA1/2020 virus and a recombinant (r)SARS-CoV-2 virus that is identical to USA-WA1/2020 except for the E484K mutation introduced in the spike RBD. We selected 34 sera from study participants based on their SARS-CoV-2 spike ELISA antibody titer (negative [N=4] versus weak [N=8], moderate [N=11] or strong positive [N=11]). In addition, we included sera from five individuals who received two doses of the Pfizer SARS-CoV-2 vaccine BNT162b2. Serum neutralization efficiency was lower against the E484K rSARS-CoV-2 (vaccination samples: 3.4 fold; convalescent low IgG: 2.4 fold, moderate IgG: 4.2 fold and high IgG: 2.6 fold) compared to USA-WA1/2020. For some of the convalescent donor sera with low or moderate IgG against the SARS-CoV-2 spike, the drop in neutralization efficiency resulted in neutralization ID_50_ values similar to negative control samples, with low or even absence of neutralization of the E484K rSARS-CoV-2. However, human sera with high neutralization titers against the USA-WA1/2020 strain were still able to neutralize the E484K rSARS-CoV-2. Therefore, it is important to aim for the highest titers possible induced by vaccination to enhance protection against newly emerging SARS-CoV-2 variants. Two vaccine doses may be needed for induction of high antibody titers against SARS-CoV-2. Postponing the second vaccination is suggested by some public health authorities in order to provide more individuals with a primer vaccination. Our data suggests that this may leave vaccinees less protected against newly emerging variants.

## Introduction

One year in the coronavirus disease 2019 (COVID-19) pandemic, the first vaccines are being rolled out under emergency use authorizations. Recently, rapidly spreading variants of severe acute respiratory syndrome coronavirus 2 (SARS-CoV-2), the virus that causes COVID-19, have been reported. It is of great concern that these newly emerging variants can escape neutralizing antibodies induced by previous infection and/or vaccination through mutations in the spike (S) protein, including the receptor binding domain (RBD), a target for neutralizing antibodies. We and others have previously reported that the asparagine (N) to tyrosine (Y) substitution at position 501 (N501Y), present in variants of concern belonging to the B.1.1.7, B.1.351 and P.1 lineages, does not seem to affect *in vitro* neutralization of SARS-CoV-2 viruses by human sera from convalescent or vaccinated human donors. However, there remains concern about additional substitutions like E484K present in B.1.351 and P.1 lineages allowing escape from neutralizing antibodies (1–4), thereby potentially rendering vaccine-induced immunity less protective.

In order to investigate the impact of the E484K mutation in the neutralizing activity of SARS-CoV-2 specific antisera, we performed *in vitro* microneutralization assays with both the USA-WA1/2020 virus and a recombinant (r)SARS-CoV-2 virus that is identical to USA-WA1/2020 except for the E484K mutation introduced in the spike RBD.

The E484K mutant rSARS-CoV-2 was generated using previously described reverse genetics based on the use of a bacterial artificial chromosome (BAC) (5–7). The USA-WA1/2020 reflects SARS-CoV-2 strains that circulated in the early phase of the COVID-19 pandemic. A total of 34 sera were selected from study participants based on their SARS-CoV-2 S enzyme linked immunosorbent assay (ELISA) antibody titer (negative [N=4] versus weak [N=8], moderate [N=11] or strong positive [N=11]). In addition, we included sera from five individuals who received two doses of the Pfizer SARS-CoV-2 vaccine BNT162b2 (V1-V5). Demographics and available metadata for each participant is summarized in Supplementary Table 1. We performed all experiments in a blinded manner. The same sera have been tested for neutralization studies with a N501Y SARS-CoV-2 variant in our recent report (8).

## Results

Sera from vaccinated donors gave high neutralization titers, similar to convalescent samples with the highest neutralization titers. Serum neutralization efficiency was lower against the E484K rSARS-CoV-2 (vaccination samples: 3.4 fold; convalescent low IgG: 2.4 fold, moderate IgG: 4.2 fold and high IgG: 2.6 fold based on geometric means) which was significantly different for the convalescent sera (see Figure 1), suggesting that the single E484K mutation in the RBD affects binding by serum polyclonal neutralizing antibodies from both convalescent and vaccinated donors. In the case of convalescent donor sera with low or moderate IgG against SARS-CoV-2 S protein, the drop in neutralization efficiency could result in neutralization ID_50_ values similar to negative control samples, resulting in low or even absence of neutralization of the E484K recombinant virus by those sera.

**Figure 1.**
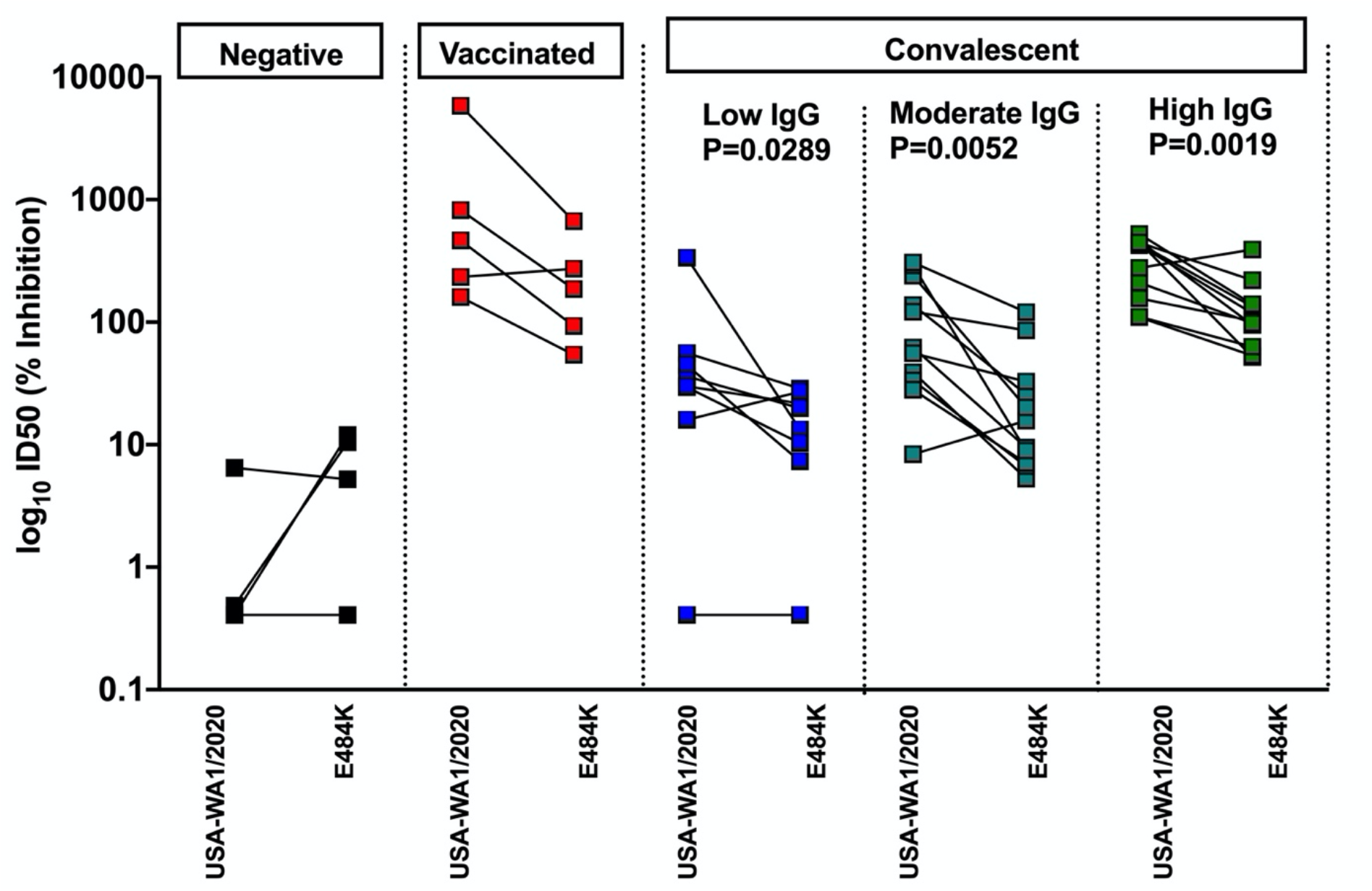
Human convalescent and post-vaccination sera neutralize E484K rSARS-CoV-2 less efficient than USA-WA1/2020 in an in vitro microneutralization assay. Convalescent sera are subdivided in low, moderate and high IgG classes based on anti-spike ELISA titers. Two-sided Mann Whitney-U tests were performed to calculate statistical differences.

## Conclusions

These data indicate that the E484K mutation present in circulating SARS-CoV-2 strains that belong to the B.1.351 and P.1 lineages reduces the neutralizing activity of human polyclonal sera induced in convalescent (infected with previous strains) and vaccinated individuals. The significant impact of a single point mutation in the neutralizing activity of polyclonal sera highlights the need for the rapid characterization of SARS-CoV-2 variants. However, human sera with high neutralization titers against the USA-WA1/2020 strain were still able to neutralize the E484K rSARS-CoV-2. Therefore, it is important to aim for the highest titers possible induced by vaccination, as this should enhance the chances for protection even in the case of antigenic drift of circulating SARS-CoV-2 strains. Currently deployed SARS-CoV-2 vaccines are recommended as a prime-boost regimen. Because of vaccine shortage and relatively strong seroconversion being observed after a single dose, some public health authorities recommended to postpone the second booster vaccination in order to be able to provide more individuals with a first primer vaccination. This will result in lower neutralizing antibody titers. Our data show that this may be problematic in the context of newly emerging SARS-CoV-2 variants, as it may leave some vaccinees unprotected. It is currently unknown which neutralization titer correlates with (full) protection, and to what extent immune mechanisms beyond direct virus neutralization contribute to protection, especially for specific target groups with comorbidities that are currently being prioritized for vaccination.

Viruses that belong to the B.1.351 and P.1 lineages have originally been described in the Republic of South Africa and Brazil, but are now reported on multiple continents already. Therefore, while it is premature to update vaccines based on these lineages, it is important that the worldwide vaccination effort will aim at fully vaccinating as many people as possible using vaccination strategies that result in induction of high neutralizing antibody titers.

## Data Availability

All data will be available upon request from the corresponding authors.

## Acknowledgements

PVI study group (in alphabetical order): H. Alshammary, A. Amoako, M. Awawda, K. Beach, C. M. Bermúdez-González, R. Chernet, L. Eaker, E. Ferreri, D. Floda, C. Gleason, Dr. G. Kleiner, D. Jurczyszak, J. Matthews, W. Mendez, Dr. LCF Mulder, K. Russo, A. Salimbangon, Dr. M. Saksena, A.Shin, L. Sominsky and K. Srivastava.

We thank all the study participants for their continued support of COVID19 research. This research was partly funded by CRIP (Center for Research for Influenza Pathogenesis), a NIAID supported Center of Excellence for Influenza Research and Surveillance (CEIRS, contract # HHSN272201400008C); by NCI grant U54CA260560; by the generous support of the JPB Foundation and the Open Philanthropy Project (research grant 2020-215611 (5384); and by anonymous donors to A.G-S. Work on SARS-CoV-2 in the Krammer and Simon laboratories was funded by the Collaborative Influenza Vaccine Innovation Centers (CIVIC) contract 75N93019C00051. Research in Martinez-Sobrido laboratory was partially funded by the New York Influenza Center of Excellence (NYICE), a member of the National Institute of Allergy and Infectious Diseases (NIAID), National Institutes of Health (NIH), Department of Health and Human Services, Centers of Excellence for Influenza Research and Surveillance (CEIRS) contract No. HHSN272201400005C (NYICE).

## Supplementary Methods section

### 50% tissue culture infective dose (TCID_50_) calculation and *in vitro* microneutralization assay

To estimate the neutralizing efficiency of human sera, *in vitro* microneutralization assays were performed. Human sera were inactivated at 56°C for 30 min. Serum samples were serially diluted 3-fold starting from 1:30 dilution in Vero-E6-infection medium (DMEM+ 2% FBS+ 1% non-essential amino acids). The samples were incubated with 450 tissue culture infective dose 50 (TCID_50_) of either USA-WA1/2020 or rSARS-CoV-2 E484K for 1 hour in an incubator at 37°C, 5% CO_2_ followed by incubation with pre-seeded Vero-E6 at 37°C for 48 hours. The plates were fixed in 4% formaldehyde at 4°C overnight. For TCID_50_ calculation, the virus stock was serially diluted 10-fold starting with 1:10 dilution and incubated on Vero-E6 cells for 48 hours followed by fixation in 4% Formaldehyde. The cells were washed with 1xPBS and permeabilized with 0.1% Triton X-100 in 1XPBS. The cells were washed again and blocked in 5% non-fat milk in 1xPBS+ 0.1% Tween-20 for 1 hour at room temperature. After blocking, the cells were incubated with anti-SARS-CoV-2 NP and anti-spike monoclonal antibodies, mixed in 1:1 ratio, for 1.5 hours at room temperature. The cells were washed in 1xPBS and incubated with 1:5000 diluted HRP-conjugated anti-mouse IgG secondary antibody for 1 hour at RT followed by a brief PBS wash. Finally, 100μl tetramethyl benzidine (TMB) substrate was added and incubated at RT until blue color appeared, and the reaction was terminated with 50μl 1M H_2_SO_4_. Absorbance was recorded at 450nm and 650nm and percentage reduction in infection was calculated as compared to negative control.

**Supplementary Table 1:**
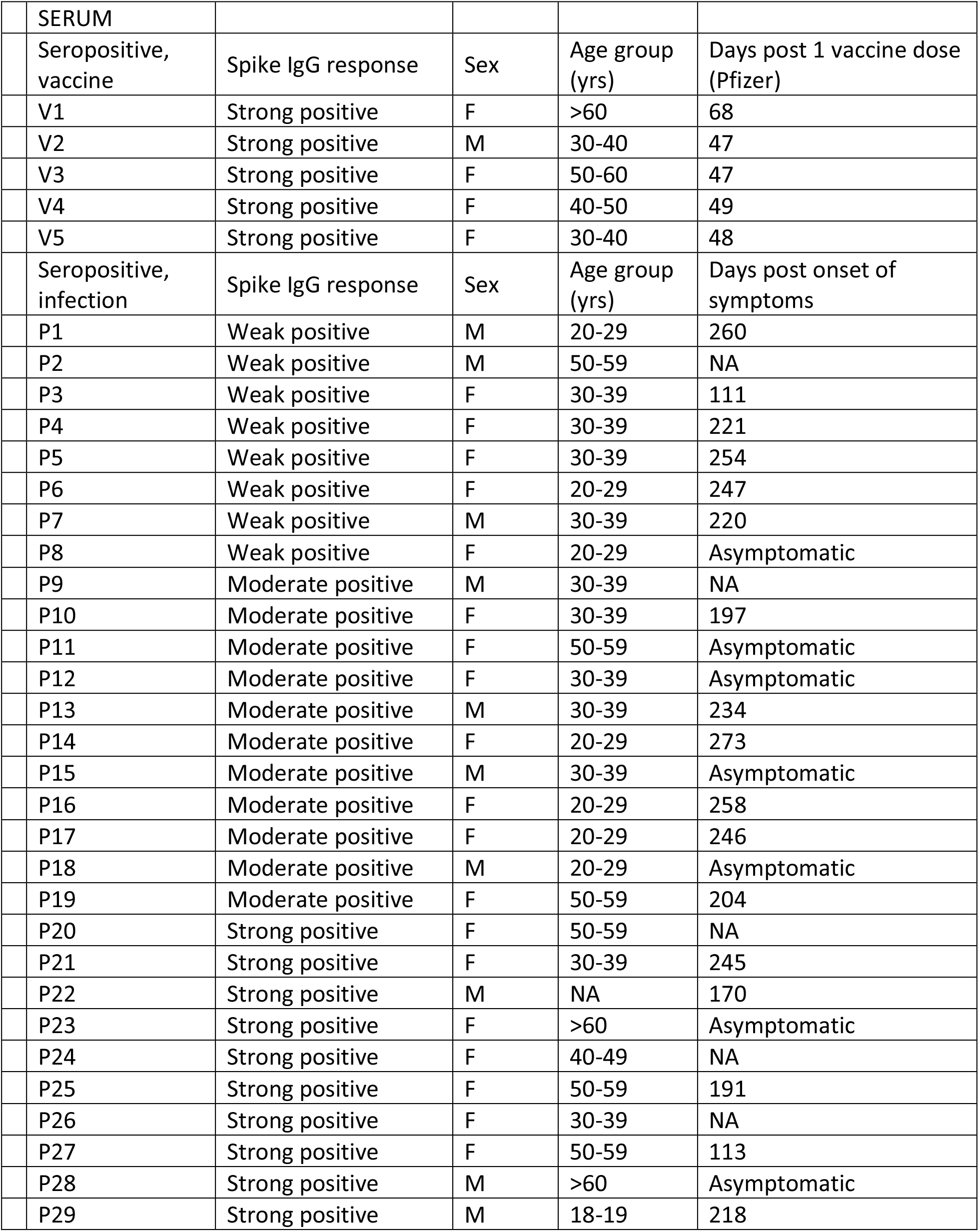
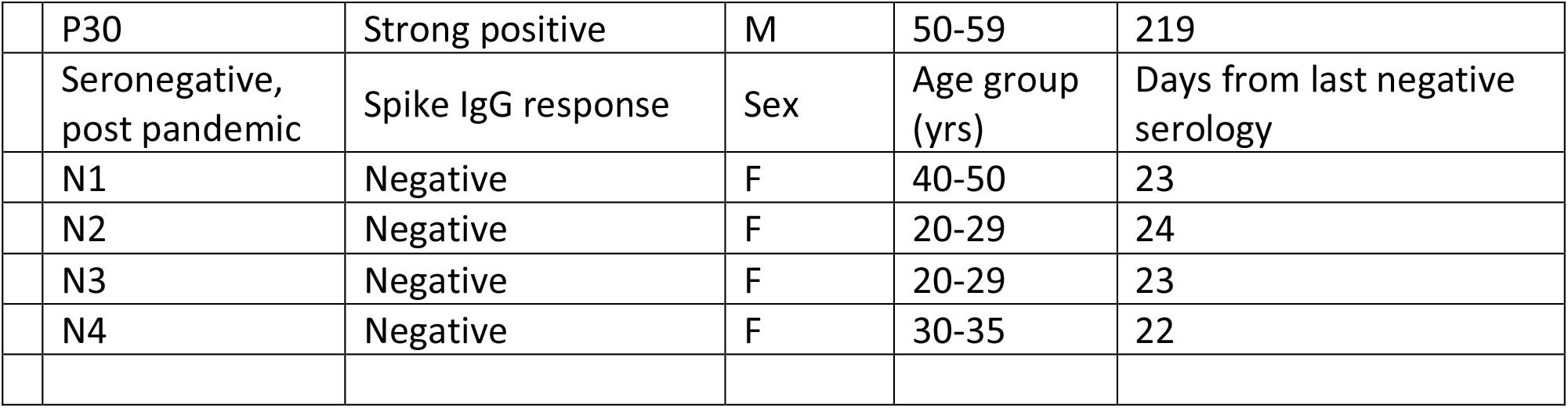
Description of serum samples obtained from human subjects.

| SERUM |  |  |  |  |
| --- | --- | --- | --- | --- |
| Seropositive, vaccine | Spike IgG response | Sex | Age group (yrs) | Days post 1 vaccine dose (Pfizer) |
| V1 | Strong positive | F | >60 | 68 |
| V2 | Strong positive | M | 30-40 | 47 |
| V3 | Strong positive | F | 50-60 | 47 |
| V4 | Strong positive | F | 40-50 | 49 |
| V5 | Strong positive | F | 30-40 | 48 |
| Seropositive, infection | Spike IgG response | Sex | Age group (yrs) | Days post onset of symptoms |
| P1 | Weak positive | M | 20-29 | 260 |
| P2 | Weak positive | M | 50-59 | NA |
| P3 | Weak positive | F | 30-39 | 111 |
| P4 | Weak positive | F | 30-39 | 221 |
| P5 | Weak positive | F | 30-39 | 254 |
| P6 | Weak positive | F | 20-29 | 247 |
| P7 | Weak positive | M | 30-39 | 220 |
| P8 | Weak positive | F | 20-29 | Asymptomatic |
| P9 | Moderate positive | M | 30-39 | NA |
| P10 | Moderate positive | F | 30-39 | 197 |
| P11 | Moderate positive | F | 50-59 | Asymptomatic |
| P12 | Moderate positive | F | 30-39 | Asymptomatic |
| P13 | Moderate positive | M | 30-39 | 234 |
| P14 | Moderate positive | F | 20-29 | 273 |
| P15 | Moderate positive | M | 30-39 | Asymptomatic |
| P16 | Moderate positive | F | 20-29 | 258 |
| P17 | Moderate positive | F | 20-29 | 246 |
| P18 | Moderate positive | M | 20-29 | Asymptomatic |
| P19 | Moderate positive | F | 50-59 | 204 |
| P20 | Strong positive | F | 50-59 | NA |
| P21 | Strong positive | F | 30-39 | 245 |
| P22 | Strong positive | M | NA | 170 |
| P23 | Strong positive | F | >60 | Asymptomatic |
| P24 | Strong positive | F | 40-49 | NA |
| P25 | Strong positive | F | 50-59 | 191 |
| P26 | Strong positive | F | 30-39 | NA |
| P27 | Strong positive | F | 50-59 | 113 |
| P28 | Strong positive | M | >60 | Asymptomatic |
| P29 | Strong positive | M | 18-19 | 218 |

|  |  |  |  |  |  |
| --- | --- | --- | --- | --- | --- |
|  | P30 | Strong positive | M | 50-59 | 219 |
|  | Seronegative,<br>post pandemic | Spike IgG response | Sex | Age group<br>(yrs) | Days from last negative<br>serology |
|  | N1 | Negative | F | 40-50 | 23 |
|  | N2 | Negative | F | 20-29 | 24 |
|  | N3 | Negative | F | 20-29 | 23 |
|  | N4 | Negative | F | 30-35 | 22 |

### Serum samples from human subjects

The study protocols for the collection of clinical specimens from individuals with and without SARS-CoV-2 infection by the Personalized Virology Initiative were reviewed and approved by the Mount Sinai Hospital Institutional Review Board (IRB-16-00791; IRB-20-03374). All participants provided informed consent prior to collection of specimen and clinical information. All specimens were coded prior to processing.

